# Prediction of Left Atrial Volume Parameters from Resting ECGs and Tabular Data Using Deep Learning in the UK Biobank

**DOI:** 10.64898/2026.02.13.26346205

**Authors:** Moritz Dieing, David Brüggemann, Zareen Farukhi, Olga V. Demler

## Abstract

We present a deep learning model that predicts left atrial (LA) volume from standard 12-lead ECG recordings and basic patient data. This approach offers a low-cost, scalable alternative to MRI-based LA volume measurement, which remains the clinical gold standard but is often inaccessible. Our model performs regression directly on LA volume targets and leverages Shapley values to provide interpretable feature importance. Results highlight the predictive value of ECG signals and demonstrate that patient features such as weight and height contribute meaningfully to the estimation.

## Introduction

Left atrial (LA) volume is known to be a significant factor in evaluating cardiac function and an important marker for cardiovascular diseases (e.g. atrial fibrillation). Current clinical standard to determine this variable uses an algorithm based on measurements from heart MRI images. However, the limited availability of the technique as well as its high cost make it difficult to efficiently measure LA volume across the world.

Compared to MRI scans, a cheaper and globally more accessible method to extract heart information is the electrocardiogram (ECG). For a standard 12-lead ECG, ten electrodes are placed on the patient’s limbs and on the surface of the chest. This makes it possible to measure the overall magnitude of the heart’s electrical potential from twelve different angles over a fixed time interval. The output of the ECG is not a rather comprehensible image of the heart, but a series of plots, each corresponding to a different angle. As data of this form is much more challenging to work with, recent research has put a lot of effort into creating and training deep learning architectures that reliably extract valuable information from ECGs. In 2019, Attia et al.^1^ proposed a model to predict reduced ejection fraction, another risk marker for cardiovascular diseases, from ECG recordings. In 2020, Riberio et al.^2^ developed a CNN-based architecture which was able to recognize 6 types of anomalies in ECGs with high accuracy. Our study aims to evaluate whether it is feasible to estimate LA volume parameters from 12-lead ECG data by introducing a deep learning model that integrates ECG recordings with tabular clinical variables.

## Methods

Overall, this work makes the following key contributions:

- **Regression Model**: We introduce a regression model which predicts LA volume variable from ECG and tabular patient data.
- **Feature importance**: By computing *Shapley values*^*3*^, we can argue about the impact of each feature on the prediction.

The remainder of this section details the model architecture, the pre-training step and the dataset used for training and evaluation.

### Model

We adopted a successful CNN-based ECG classification architecture (Riberio et al.) for the regression task of predicting three different continuous LA volume variables. The adaptation mainly consisted of an additional fully connected layer enabling us to improve the model’s predictive capacity by not only using the latent ECG representation but also patient-specific tabular variables like height, weight, or age. As we observed an increase in performance, the latent representation output by the ECG encoder is a three-dimensional vector. We will refer to the *i*-th entry of this vector as “ECG repr. *i*”. Our final model was trained on batches of data applying an Adam optimizer^4^ with learning rate 0.001 and early stopping after nine epochs without improvement. Improvement was measured by computing the mean squared error (MSE) between model predictions and true outcomes.

### Outcome

The model outcome consists of three variables, namely:

- LA max volume
- LA min volume
- LA stroke volume

LA max volume corresponds to the maximum volume of the left atrium, typically occurring at the end of left ventricular systole. LA min volume is the smallest atrial volume, generally recorded at the end of left ventricular diastole. LA stroke volume is defined as, *LA stroke* = (*LA max* − *LA min*) and represents the change in volume of the left atrium during one cardiac cycle. The true outcomes of LA max and LA min volume for each patient were extracted from cardiac MRI (cMRI) images.

### Preprocessing

In our preprocessing step we only modified ECG recordings but did not touch any tabular data.

First, we applied a bandpass Butterworth filter to each ECG record. The cutoff frequency was set to 0.75*Hz* and 50*Hz* for the highpass and lowpass filters. Afterwards, the data was resampled from a sampling rate of 500*Hz* down to 400*Hz*. Finally, each record was either cropped or zero-padded such that each lead consisted of 4096 samples. Resampling and reshaping were done to match the required input shape of the existing ECG encoder from Riberio et al. (Ribeiro et al., 2020).

### Pre-training

To boost the performance of the model and to achieve faster convergence of the training procedure, the ECG encoder was first trained on the publicly avail-able PTB-XL database (https://physionet.org/content/ptb-xl/1.0.3/), which comprises 21799 labeled 12-lead ECG records. Unlike the LA volume targets, labels in the PTB-XL database are categorical. The resulting weights from this pre-training were then used to initialize the model for the main training phase.

## Results

### Dataset

We included all participants with ECG recordings in the UK Biobank database for training and evaluating the final model who also had LA volume parameters estimated from cMRI. cMRI was performed on the same day as ECG recordings.

A total of 31227 patients were enrolled. The cohort was randomly split into training (*n* = 24981), validation (*n* = 3123), and test (*n* = 3123) set. The mean age of all patients was 54.7 ± 7.5 and 52.4% (*n* = 16359) were female. The outcome variables had the following means (SD): LA max volume, 73.0 ± 23.0 mL; LA min volume, 29.4 ± 15.0 mL; and LA stroke volume, 43.5 ± 11.8 mL. Patient baseline characteristics are reported in more detail in Table 1.

**Table 1.** Baseline characteristics across dataset splits. Continuous variables are presented as mean (SD); categorical variables as n (%).

| 2. Variable | Overall | Training set | Validation set | Test set | P-Value |
| --- | --- | --- | --- | --- | --- |
| n | 31227 | 24981 | 3123 | 3123 |  |
| **Sex, n (%)** |  |  |  |  | 0.214 |
| Female | 16359 (52.4) | 13027 (52.1) | 1657 (53.1) | 1675 (53.6) |  |
| Male | 14868 (47.6) | 11954 (47.9) | 1466 (46.9) | 1448 (46.4) |  |
| Age, mean (SD) | 54.7 (7.5) | 54.7 (7.5) | 54.7 (7.4) | 54.6 (7.4) | 0.596 |
| Height, mean (SD) | 170.2 (9.4) | 170.2 (9.4) | 170.3 (9.5) | 170.1 (9.3) | 0.814 |
| Weight, mean (SD) | 75.9 (15.0) | 75.9 (15.0) | 76.0 (15.3) | 75.6 (14.8) | 0.549 |
| BSA, mean (SD) | 1.9 (0.2) | 1.9 (0.2) | 1.9 (0.2) | 1.9 (0.2) | 0.513 |
| **Ethnic Background, n (%)** |  |  |  |  | 0.503 |
| Asian or Asian British | 316 (1.0) | 259 (1.0) | 26 (0.8) | 31 (1.0) |  |
| Black or Black British | 201 (0.6) | 168 (0.7) | 19 (0.6) | 14 (0.4) |  |
| Chinese | 97 (0.3) | 77 (0.3) | 11 (0.4) | 9 (0.3) |  |
| Mixed | 148 (0.5) | 124 (0.5) | 14 (0.4) | 10 (0.3) |  |
| None | 10 (0.0) | 8 (0.0) | 1 (0.0) | 1 (0.0) |  |
| Other | 235 (0.8) | 185 (0.7) | 33 (1.1) | 17 (0.5) |  |
| White | 30220 (96.8) | 24160 (96.7) | 3019 (96.7) | 3041 (97.4) |  |
| LA stroke volume, mean (SD) | 43.5 (11.8) | 43.6 (11.8) | 43.4 (11.8) | 43.6 (11.6) | 0.750 |
| LA max volume, mean (SD) | 73.0 (23.0) | 73.0 (23.0) | 72.6 (22.9) | 73.2 (23.5) | 0.658 |
| LA min volume, mean (SD) | 29.4 (15.0) | 29.4 (14.9) | 29.2 (14.6) | 29.6 (15.9) | 0.692 |

### Model performance

After the early stopping was triggered (hence nine epochs without improving the MSE on the validation dataset), we considered that the training procedure converged. We recovered the model with the lowest MSE and set it as our final model. To evaluate the performance of this model, we computed the Spearman and Pearson correlation coefficients between predictions and true labels. As a baseline, we compared our model against a simple linear regressor using only tabular features.

Table 2 reports the average absolute Spearman and Pearson coefficients with corresponding p-values for our model, our model without pre-training and the benchmark. The average was computed across the three LA volume targets. Our model outperformed the simple benchmark model for the Pearson correlation, achieving a coefficient of **0.5308** (compared to 0.2743), as well as for the Spearman correlation, achieving a coefficient of **0.4643** (compared to 0.2559). In addition, both correlation coefficients are significantly lower without using the pre-training weight for the initialization.

**Table 1.** Average absolute Pearson and Spearman correlations.

| Model | Mean Pearson correlation (p-value) | Mean Spearman correlation (p-value) |
| --- | --- | --- |
| Benchmark | .31 ($2.46 \times 10^{-59}$) | .30 ($3.71 \times 10^{-52}$) |
| Our model | .53 ($1.49 \times 10^{-156}$) | .46 ($3.58 \times 10^{-151}$) |
| Our model (no pre-train) | .49 ($24.23 \times 10^{-150}$) | .45 ($1.15 \times 10^{-140}$) |

### Feature importance

To quantify the contribution of each input feature to the model’s prediction, we leverage Shapley values. The Shapley beeswarm plot in Figure 1 displays both the impact of the feature values on the model output, as well as a ranking of the features by importance for the prediction of the LA max volume target. Beeswarm plots for the other two targets show similar characteristics.

**Figure.**
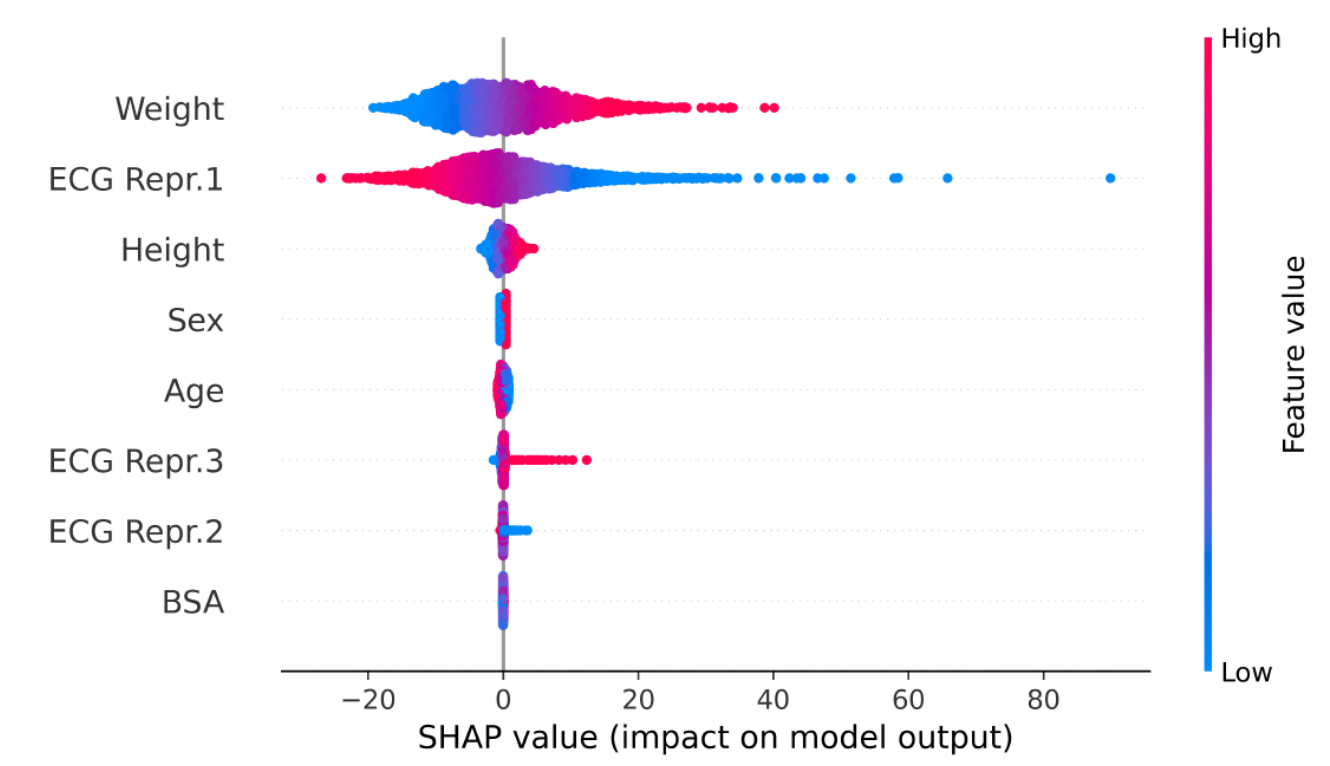
Shapley beeswarm plot for the LA max volume feature.

As the comparison between model and benchmark already suggested, the information extracted from the ECG adds value to the prediction. Weight is ranked as the most important feature. The first input of the latent ECG vector and height are the second and third most impactful features. The plot aligns with the expected behavior, showing that larger values of weight and height are associated with an increase in LA max volume. In addition, a transfer learning procedure that is not based on categorical but continuous output might lead to better initial weights.

## Conclusions

The results indicate that ECG recordings contain valuable information for predicting left atrial volume. Although the model output does not perfectly match the true labels, it achieves a significantly larger correlation coefficient than the benchmark. The results also show that the model performance benefits from the initialization with pre-trained weights.

We would like to acknowledge several limitations of our study. First, our model was trained and evaluated on a mostly White, middle-aged cohort. This may limit generalizability to more diverse populations. Second, due to the small availability of ECG and MRI quality measurements, no quality-based filtering was applied. Therefore, suboptimal input data or outcome variables may have influenced the model’s performance. Finally, our model explains variance in LA volume but does not reach perfect correlation; thus, it should be viewed as complementary rather than a replacement for the traditional MRI-based LA volume estimation. Future work could try to improve the model performance by adding more useful information through tabular variables.

## Data Availability

All data generated as part of the present study are available upon reasonable request to the authors. Access to underlying UK Biobank dataset is available after appropriate approval by UK Biobank and signing of Material Transfer Agreement/Data Use Agreement. This research has been conducted using the UK Biobank Resource under application number 206575.

https://www.ukbiobank.ac.uk/

## Acknowledgements

This research has been conducted using the UK Biobank Resource under application number 206575.

## IRB approval numbers

Business Administration System for Ethics Committees, Zurich Switzerland: BASEC-Nr.: Req-2024-00223; Brigham and Women’s Hospital Boston, MA, USA IRB protocol# 2020P002478.

